# Quality of life in patients with primary subclavian vein thrombosis, receiving early invasive treatment versus late decompressive surgery or anticoagulation alone

**DOI:** 10.1101/2025.03.19.25323973

**Authors:** Sara Lindblom, Jonas Malmstedt

## Abstract

**Introduction:** Primary subclavian vein thrombosis (PSVT) is a rare (2/100,000 per year) disorder, affecting mostly young, active and otherwise healthy individuals. Untreated it can lead to post thrombotic syndrome (PTS) which leaves the patient with a chronic impairment of the arm. Several treatment strategies have been suggested to avoid PTS: immediate thrombolysis and decompressive surgery, delayed decompressive surgery or treatment with anticoagulation alone. Studies show conflicting results, and there is scarce evidence regarding which strategy to prefer.

**Aim:** To investigate whether early invasive treatment with thrombolysis and decompressive surgery in patients with PSVT, is associated with improved quality of life compared to delayed decompressive surgery or treatment with anticoagulation alone.

**Material and methods:** Quality of life was evaluated in 87 patients treated for PSVT in Stockholm during 2008-2017. Treatment with immediate thrombolysis and surgery (acute group, n=46) was compared to treatment with delayed surgery (chronic group, n=20), and with anticoagulation alone (conservative group, n=21). The patients answered a disease-specific quality of life instrument (Disabilities of the arm, shoulder and hand, DASH) before and after intervention. One-way ANOVA and Post-hoc test was used to identify differences in DASH-score between the treatment groups. A multivariate regression model was used to adjust for potential confounders.

**Results:** The acute group had superior outcome with lower DASH-score compared to the chronic group after intervention (p=.006), and compared to the chronic group and conservative group together (p=.0002). No confounders were found.

**Conclusions:** Treatment of PSVT with immediate thrombolysis and surgery was related to superior results compared to delayed treatment. Because of the retrospective study design, more research is needed to clarify if causation is present.

## Introduction

Upper-extremity deep vein thrombosis (UEDVT) is rare and constitutes about 10% of all deep venous thrombosis (DVT) (1). The risk factors for UEDVT differ from those associated with DVT in the lower extremities. Patients with UEDVT are more likely to be younger, leaner and have no history of DVT, compared to patients who develop DVT in the lower extremities (2–5).

UEDVT is a broad category of thrombosis localized in several different vein segments in the upper extremity. UEDVT can be divided into two etiological groups: primary and secondary. Secondary UEDVT, which is the most common, has an underlying cause such as cancer or the use of central venous catheters (CVC) or pacemaker leads within the affected vein. The major risk factor to develop secondary UEDVT is the use of CVC, which increases the odds seven-fold (2). Primary UEDVT has no underlying disease or known trigger factor.

### Terminology

Primary subclavian vein thrombosis (PSVT) is a separate entity with an underlying anatomic abnormality causing compression of the subclavian vein within the thoracic outlet. The anterior thoracic outlet is the space in between the clavicle, first rib and anterior scalene muscle. The presence of abnormal anatomy or hypertrophy of the muscles in this area narrows the outlet and the surrounding structures compress the subclavian vein. This condition is also known as venous thoracic outlet syndrome (V-TOS) (6).

PSVT, a rare disorder with an yearly incidence of approximately 2/100,000 inhabitants (6), consists of idiopathic and effort-related causes. Idiopathic cases have no known cause or trigger factor whilst the effort related cases, also called Paget-Schroetter syndrome, is triggered by strenuous activities of the arm (7).

### Pathogenesis

PSVT usually presents with a sudden onset of a swollen, painful, heavy and blue arm. The presence of superficial dilated veins on the shoulder, chest and neck is common. PSVT is more common in young otherwise healthy individuals in their early thirties (6).

The pathogenesis of PSVT is thought to be frequent micro-trauma to the subclavian vein at the passage between the clavicle, first rib and anterior scalene muscle due to strenuous work with the arm in an elevated position. During strenuous elevated work (eg. badminton, painting and auto repair), the vein is pinched in between the clavicle and the first rib, which causes repeated micro-trauma. The micro-trauma causes damage to the vein and induces a local inflammation, which in turn results in fixation of the vein to surrounding structures, fibrosis of the vein wall and a thickening of the intima of the vessel. The vein lumen becomes narrower which reduces the blood flow, and occlusion can occur over time (8).

### Complications

The risk of developing pulmonary embolism (PE) is present for all patients with DVT. Compared to DVT in the lower extremities and secondary UEDVT, PSVT has a lower risk to develop PE. Kooij et al showed that the incidence of PE in patients with secondary UEDVT was twice as high as for patients with PSVT (9).

Post-thrombotic syndrome (PTS) is a chronic complication due to venous thrombosis characterized by swelling, oedema, pain and heaviness in the affected limb (10). There is limited data on PTS following UEDVT, and there is no gold standard for diagnosis of PTS in the upper extremities (11). The studies available report an incidence of 6-44% for PTS following UEDVT (12, 13), but since the criteria for PTS in the upper-extremity is not coherent and different aetiologies (primary and secondary) for UEDVT were included in the studies, the true incidence for PTS following PSVT is uncertain. There seems to be a difference in the development of PTS between secondary and primary UEDVT where CVC-induced UEDVT has a lower risk for development of PTS whilst PSVT has a higher risk of developing PTS (11, 14).

### Quality of life instruments

In order to evaluate the symptoms and physical function in patients with PSVT, different instruments can be used. “The Disabilities of the Arm, Shoulder and Hand” (DASH) is a frequent used scoring-system where the patient answers 30 questions regarding symptoms and function of the affected limb. The DASH-score is then calculated and converted in to a scale from 0 (no symptoms) to 100 (worst symptoms) points. In a normal population, the DASH-score is age-dependent with higher score the older the population (15). The DASH-score has proven to perform well in describing symptoms and limitations of the upper extremities and monitoring changes over time (16, 17). The Swedish version of the DASH-instrument has been proven to be a reliable measurement of upper extremity disabilities (18).

A small pilot-study of quality of life (QoL) after PTS performed by Khan et al suggested that the development of PTS affects the QoL in patients with UEDVT (13). The quality of life was measured with DASH-score and was compared between patients with and without PTS. This is important since PTS is common also after PSVT, primarily affecting young people with high expectations of normal arm function. The need to prevent PTS in order to prevent a decline in QoL in this patient group is therefore desirable.

### Treatment

The purpose of the treatment for PSVT is to relief the symptoms of the thrombosis, prevent recurrent thrombosis and PE, and to prevent PTS. Treatment guidelines are based on data from retrospective studies and clinical experience, as there are no randomized controlled trials of the treatment for PSVT (19).

The conventional treatment of PSVT consists of anticoagulation, primarily to prevent PE and thrombus extension. The general guideline is the use of vitamin K antagonist (VKA) for >3 months after thrombosis (19), but the use of anticoagulation alone is often not enough to prevent residual thrombosis and remaining symptoms. In a small retrospective study, Persson et al showed that 77% of patients with PSVT treated with anticoagulation alone had remaining symptoms in the affected arm, and 55% had residual thrombosis (20). Similar results were shown in a retrospective study performed by Urschel et al (21).

Catheter-directed thrombolysis (CDT) is an alternative first step in the treatment of PSVT. The success rate is dependent on the timing of the thrombolysis. If thrombolysis is performed within 14 days from onset of symptoms, a complete thrombolysis can be achieved in 62-84% of the patients, where the earlier thrombolysis the better (22–24). Thrombolysis performed later than 14 days from symptom onset is rarely successful. Urschel et al showed that when thrombolysis was performed after 6 weeks from the time of venous occlusion, a decrease of the thrombus was only seen in 50% of the patients, and none was completely resolved (25). Although thrombolysis can dissolve the thrombosis, it does not prevent recurrent thrombosis. The cause of the thrombosis is still present, and recurrence is frequent. Even if thrombolysis is successful initially, the incidence of recurrent symptoms is high in patients only receiving catheter-directed thrombolysis (25, 26).

In order to treat the underlying cause of the thrombosis in patients with PSVT, an intervention with decompressive surgery of the thoracic outlet in addition to CDT is now advocated by some centres (21, 27–29). During the surgical procedure, the anterior part of the first rib is removed and fibrotic tissue surrounding the affected vein is excised (venolysis). There have been promising results regarding vein patency and subjective symptom relief following this invasive strategy (21, 27, 29). Urschel and colleagues showed good results for patients treated with surgery compared to those treated with anticoagulation alone (21, 25).

There are several surgical approaches but no evidence on which approach is to be preferred. The transaxillary approach is most common, but may be technically challenging and associated with a higher risk for complications such as injury to peripheral nerves. Alternative approaches are supra-, para and infraclavicular first rib resection. Whatever method is used, it is important that the first rib is resected and that the fibrotic tissue surrounding the vein is removed (6).

The placement of stents within the subclavian vein is not recommended for PSVT (19) because fracture and deformation of the stent, and rethrombosis is common (30). The costoclavicular junction initiates a force that easily can deform the stent and the wide range of movement in the shoulder can make the inflexible stent perforate the vein.

### Treatment strategies

Since there are no randomized clinical trials for the treatment of PSVT, there is no level I evidence pointing to which strategy is superior. It is also not known what the true incidence of PTS is for PSVT, and if invasive treatment is needed in all cases. The treatment strategies used can be divided into three groups: conventional treatment with anticoagulation alone, early thrombolysis combined with early decompressive surgery and conventional treatment with delayed surgery in case the patient search medical assistance too late for thrombolysis to be performed. As described above, treatment with anticoagulation alone seems not to be optimal because of the high incidence of remaining symptoms and recurrent thrombosis.

The combination of early thrombolysis and decompressive surgery has shown to give promising results. In a large study by Urschel et al, 96% of patients treated with thrombolysis and surgery were regarded having good to excellent results (21). This was the first large series regarding treatment for PSVT, but only 25 patients were treated according to modern regime with early CDT and surgery. Furthermore, the patients were not evaluated with QoL-forms but only with three arbitrary factors (pain relief, employment, limited recreation).

Another study performed by Molina et al had a 100% patency of the vein after early thrombolysis and prompt surgery. They had a different approach to the surgery were they also reconstructed the damaged vein with a patch. This study only examined vein patency and not how the results affected the everyday life or QoL (27).

One recent study by Taylor et al, suggested that long-term outcome (measured with the short DASH-score) was improved by surgery but not affected if thrombolysis was performed or not before delayed surgery. The study also suggested that chronic rethrombosis of the subclavian vein did not result in poorer outcome (also measured with DASH-score) compared to patients with patency of the affected vein. This was discussed to be a result of the formation of collateral veins in patients with a chronic occlusion of the vein (29). However, this study had some limitations making it difficult to interpret the results. The results described above were not proven statistically significant. The study was performed at a referral centre and the patients referred and treated with delayed surgery alone might have been the patients with the most severe symptoms, whilst patients with less severe symptoms perhaps were not referred at all. This makes it a selected population and the results cannot be generalized.

Molina et al showed that delayed surgery in a selected group of patients with a short segment of thrombosis did provide a 100% patency of the affected vein (27). Limitations to this study were that QoL was not described for the patients and only a selected group of patients were treated with delayed surgery.

Guzzo with colleagues had similar results for vein patency in the group treated with anticoagulation alone before delayed decompressive surgery compared to the group treated with thrombolysis prior to delayed surgery. Overall 91% had patent veins at follow up, and all had a relief of symptoms (31).

Since there are conflicting results and scarce evidence regarding which strategy to use for PSVT, more research is needed to provide information of how this patient group preferably should be treated in order to minimize PTS and to improve QoL. With this study we hope to bring some clarity to what treatment strategy is to be favored for PSVT patients.

### Aim

The aim was to investigate whether early invasive treatment with thrombolysis and decompressive surgery in patients with primary subclavian vein thrombosis, is associated with improved quality of life compared to delayed decompressive surgery or treatment with anticoagulation alone.

## Materials and Methods

This study was a cohort study of patients in the county of Stockholm treated for PSVT at Södersjukhuset and the Karolinska university hospital. This is part of Sara Lindbloms Degree project 30 credits, spring 2017 at the Study Program in Medicine at Karolinska Institutet.

### Study Population

The study population consisted of 87 patients treated for PSVT with anticoagulation alone or early or late invasive treatment during the past 9 years. Early invasive treatment was commenced in Stockholm in 2010 and was the reason why data was not available from earlier years.

The study population was identified through the quality registry for vascular surgery (Swedvasc) and the inclusion criteria of this study were patients diagnosed with PSVT and treated at Södersjukhuset or Karolinska university hospital. The exclusion criteria and exclusion process can be seen in Figure I. To be noted is that patients developing PSVT during pregnancy were excluded from the study due to that active treatment could not be given irrespectively of timing of the diagnosis of PSVT.

**Figure I.**
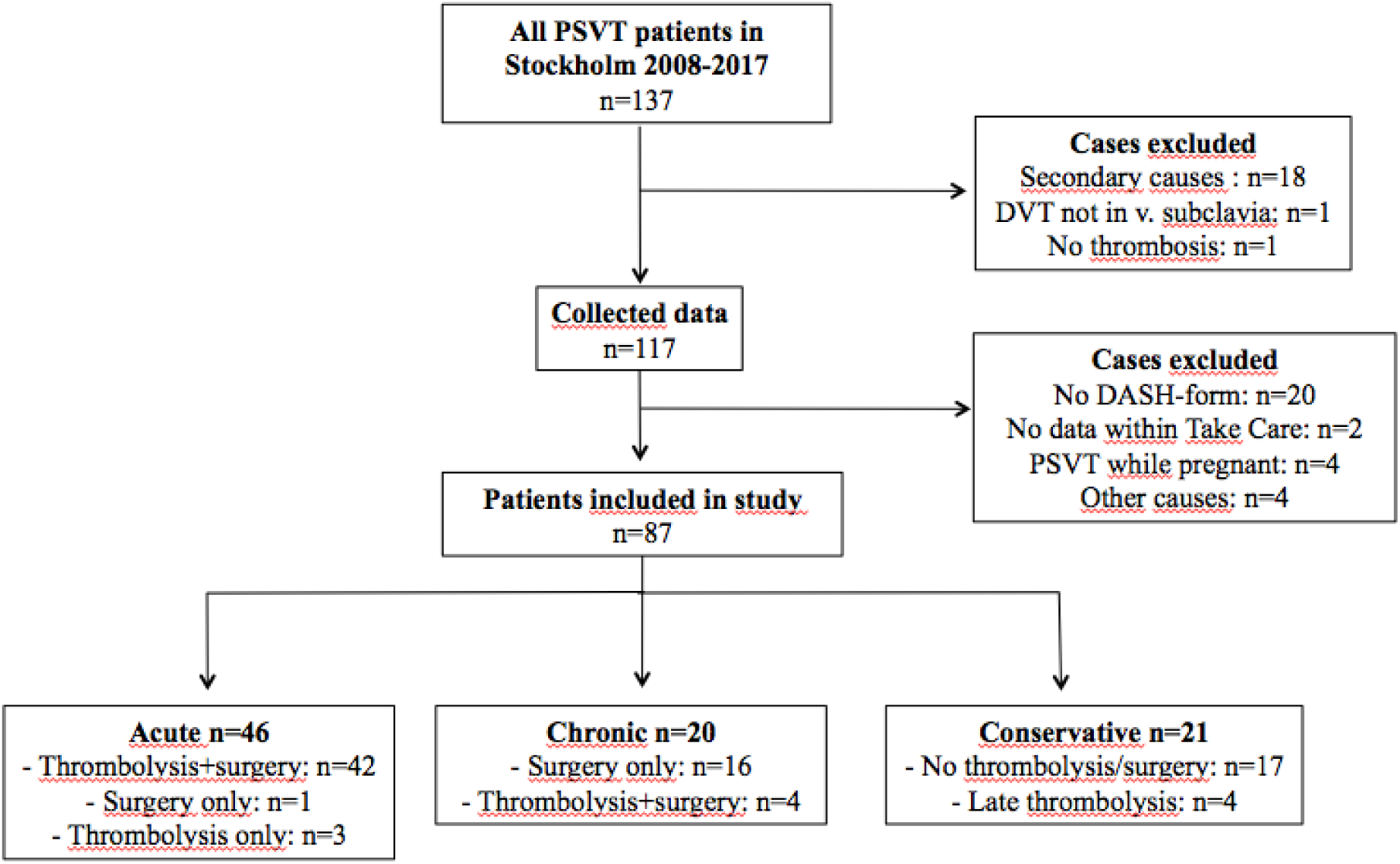
Flow-chart displaying patient selection. Secondary causes consisted of: thrombosis due to the use of CVC/pacemaker-/ICD-leads within the affected vein (*n*=7), clavicle fracture (*n*=5), malignancy (*n*=4), skin infection (*n*=1) and Leishmaniasis (*n*=1). Other causes for exclusion of patients were: not treated in Sweden (*n*=2), did not speak Swedish (*n*=1) or were too young of age (*n*=1).

### Diagnosis and choice of treatment

Most patients within the study population were diagnosed with ultrasonography, which have a sensitivity of 97% and a specificity of 96% for diagnosing UEDVT (32). In case of inconclusive ultrasonography findings, a venography was performed and if patient had symptoms of PE, a CT-scan was performed.

Symptomatic patients that were diagnosed within two weeks from thrombosis were offered immediate thrombolysis and surgery. Patients that were diagnosed or referred to vascular surgeon later than two weeks from thrombosis were initially treated with anticoagulation. If the patients in this group experienced severe and persisting symptoms, surgery was offered if symptom relief was thought to be achievable. Patients that refused surgery, had a thrombosis earlier than 2010 or were not believed to benefit from surgery were treated with anticoagulation alone.

The study population was divided into three groups depending on timing and type of treatment (Figure I). The acute group consisted of patients treated with early thrombolysis and surgery, three patients that received only early thrombolysis and one patient that received only early surgery. The chronic group consisted of patients that received late surgery only and four patients that received both late thrombolysis and surgery. The conservative group consisted of patients treated with anticoagulation alone, and four patients that received late thrombolysis that was not successful (no regression of the thrombus and no improvement of the acute symptoms) i.e. could be accounted for as conservatively treated.

Thrombolysis was performed as local catheter-directed thrombolysis with recombinant tissue plasmine activator (Actilyse®). Control of the thrombolysis-process was made daily and thrombolysis was terminated when complete resolution of thrombus was achieved. The thrombolysis was also stopped if there was no reduction of remaining thrombus between two controls.

All the patients that received surgery were operated using the infraclavicular approach, where the skin incision is placed inferior to the clavicle. The anterior part of the first rib was resected in order to relieve the chronic compression of the vein. Fibrotic tissue surrounding the vein was excised in order to remove the extrinsic component of the stenosis. After the surgery, patients received rehabilitation led by a physiotherapist.

### Data collection

Patients treated for PSVT during the past 9 years in Stockholm have answered QoL-surveys and been examined with duplex ultrasonography in order to support the clinical decisions during treatment. The data has been collected before the surgery, one month after surgery, and one year after surgery. The QoL-survey used was the DASH-score and the forms were either answered during consultation at the hospital or sent by mail to the patient before medical appointment. New QoL surveys were also sent to all patients enrolled in the study in order to get a longer follow-up. The collected QoL-forms were divided into groups: DASH-forms answered at time of thrombosis were considered to be DASH-score baseline, DASH-forms answered <100 days from thrombosis were considered short-term and DASH-forms answered >100 days from thrombosis were considered long-term.

Patient demographics, such as age, gender, handedness and occupation were collected from patient medical records. Risk factors such as strenuous activities adjacent to the thrombosis, known clotting disorder and the use of oral contraceptives were collected for each patient. The data from the QoL forms and medical records was entered in to EpiData software, version 4.0.2.49 (Comprehensive Data Management and Basic Statistical Analysis System. Odense. Denmark). The data was coded and only personnel involved in the study had the access to the data.

### Primary outcome

The primary outcome was the difference in long-term disease-specific QoL(measured by DASH-score) between the three different treatment stretegies. Long-term QoL in patients treated with early invasive treatment was compared to long-term QoL in patients treated with late invasive treatment, and patients treated with anticoagulation alone.

### Statistical analysis

Descriptive statistics was used for baseline characteristics. Continuous variables with normally distributed data were described with mean and 95% confidence intervals (CI), and median and inter-quartile range (IQR) was used for continuous variables with non-normal distribution. Categorical variables were described with proportions and 95% CI for proportions (using the Wilson score method).

Comparison between the groups regarding baseline, short-term and long-term DASH-score was performed using One-Way ANOVA. To identify between which groups there was a significant difference, a Games-Howell Post Hoc test was used because of the differences in sample size and variance between the groups.

A multivariable linear regression model was used to discover and adjust for potential covariates affecting long-term DASH-score. Age, sex, smoking and whether the dominant arm was affected, was taken in to account in the multivariate analysis.

The analyses were performed using SPSS version 24 (IBM, Armonk, NY, USA) and CI in Table I was calculated with the online tool OpenEpi version 3.01 (33). The minimum clinically important difference in DASH-score before versus after intervention was set to 13 points as advised in the third edition of the DASH-manual (34). A p-value <.05 was regarded as significant.

**Table I.**
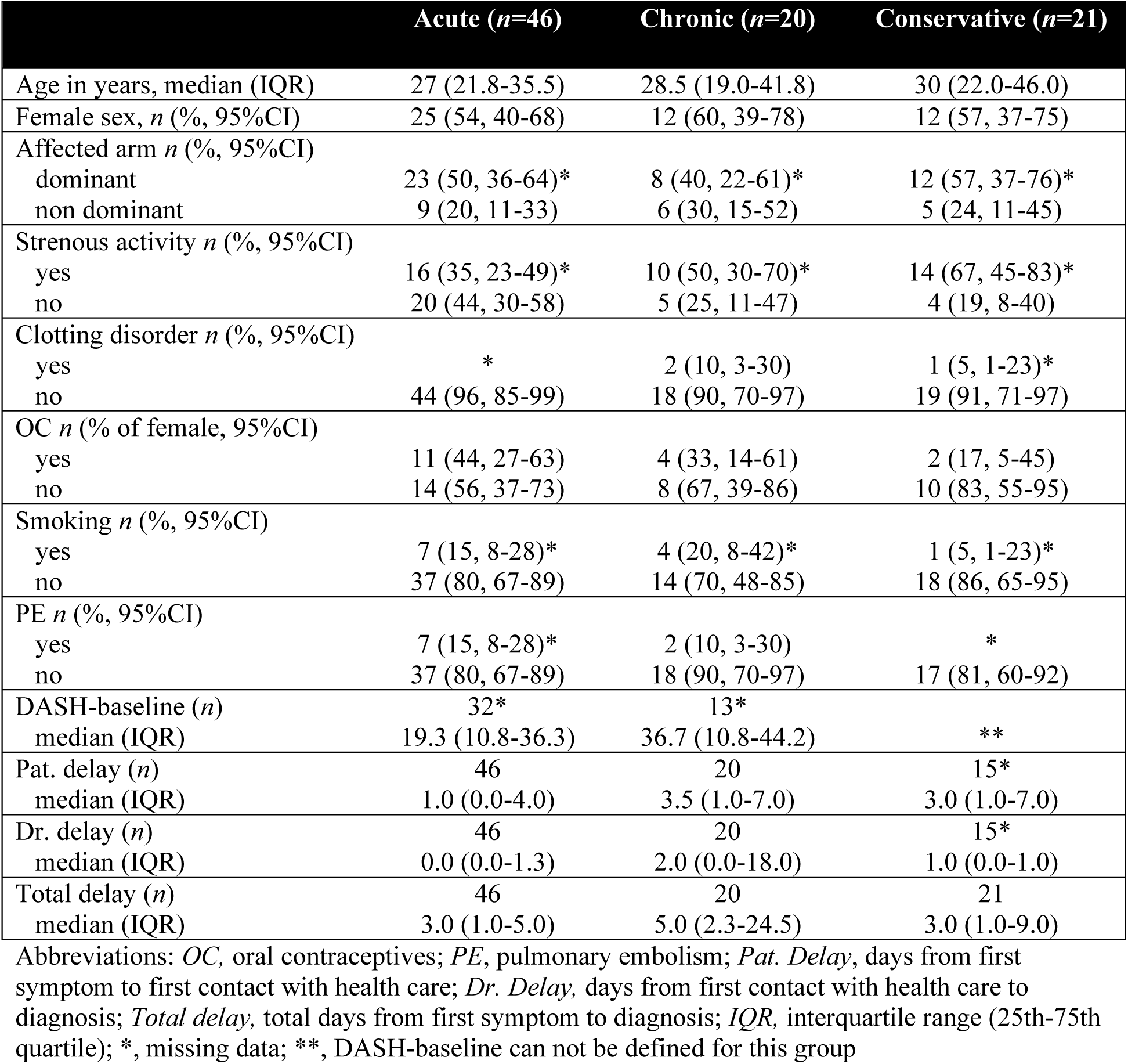
Baseline characteristics at time of thrombosis. Characteristics of the study population are presented for the 3 treatment groups: early thrombolysis (within 2 weeks) and surgery (acute), delayed surgery (chronic) and non-surgical treatment (conservative).

|  | Acute (n=46) | Chronic (n=20) | Conservative (n=21) |
| --- | --- | --- | --- |
| Age in years, median (IQR) | 27 (21.8-35.5) | 28.5 (19.0-41.8) | 30 (22.0-46.0) |
| Female sex, n (% , 95%CI) | 25 (54, 40-68) | 12 (60, 39-78) | 12 (57, 37-75) |
| Affected arm n (% , 95%CI) |  |  |  |
| dominant | 23 (50, 36-64)* | 8 (40, 22-61)* | 12 (57, 37-76)* |
| non dominant | 9 (20, 11-33) | 6 (30, 15-52) | 5 (24, 11-45) |
| Strenous activity n (% , 95%CI) |  |  |  |
| yes | 16 (35, 23-49)* | 10 (50, 30-70)* | 14 (67, 45-83)* |
| no | 20 (44, 30-58) | 5 (25, 11-47) | 4 (19, 8-40) |
| Clotting disorder n (% , 95%CI) |  |  |  |
| yes | * | 2 (10, 3-30) | 1 (5, 1-23)* |
| no | 44 (96, 85-99) | 18 (90, 70-97) | 19 (91, 71-97) |
| OC n (% of female, 95%CI) |  |  |  |
| yes | 11 (44, 27-63) | 4 (33, 14-61) | 2 (17, 5-45) |
| no | 14 (56, 37-73) | 8 (67, 39-86) | 10 (83, 55-95) |
| Smoking n (% , 95%CI) |  |  |  |
| yes | 7 (15, 8-28)* | 4 (20, 8-42)* | 1 (5, 1-23)* |
| no | 37 (80, 67-89) | 14 (70, 48-85) | 18 (86, 65-95) |
| PE n (% , 95%CI) |  |  |  |
| yes | 7 (15, 8-28)* | 2 (10, 3-30) | * |
| no | 37 (80, 67-89) | 18 (90, 70-97) | 17 (81, 60-92) |
| DASH-baseline (n) | 32* | 13* |  |
| median (IQR) | 19.3 (10.8-36.3) | 36.7 (10.8-44.2) | ** |
| Pat. delay (n) | 46 | 20 | 15* |
| median (IQR) | 1.0 (0.0-4.0) | 3.5 (1.0-7.0) | 3.0 (1.0-7.0) |
| Dr. delay (n) | 46 | 20 | 15* |
| median (IQR) | 0.0 (0.0-1.3) | 2.0 (0.0-18.0) | 1.0 (0.0-1.0) |
| Total delay (n) | 46 | 20 | 21 |
| median (IQR) | 3.0 (1.0-5.0) | 5.0 (2.3-24.5) | 3.0 (1.0-9.0) |

Abbreviations: *OC*, oral contraceptives; *PE*, pulmonary embolism; *Pat. Delay*, days from first symptom to first contact with health care; *Dr. Delay*, days from first contact with health care to diagnosis; *Total delay*, total days from first symptom to diagnosis; *IQR*, interquartile range (25th-75th quartile); \*, missing data; \*\*, DASH-baseline can not be defined for this group

### Ethical considerations

Since this study consisted of clinical data in already treated patients, there was no risk for physical harm regarding the patients enrolled in the study. The ethical issue of this study was that the reading of medical records and the dispatch of QoL forms might be seen as an intrusion. Patients were not asked for permission to read their medical records, but they were informed about the implementation of this study when mailed the QoL forms. In the mailed forms it was made clear that the participation was non-compulsory and that the patient could decide to whether fill in or not fill in the questionnaire. Patients who did not understand Swedish were not asked to respond to the QoL forms.

The data used in this study was registered in case report forms (CRFs). The information in the CRFs was coded and did not contain names or personal number of the enrolled patients. The list of codes to decrypt the data was locked up separately from the CRF forms. Only researchers involved in the study had the access to the CRF forms and the key to unlock the data. The CRFs will be stored at Södersjukhuset for 10 years and thereafter destroyed.

Our valuation was that the advantages of this study (to learn what clinical strategy is the best for PSVT) outbalanced the intrusion in privacy for the participating patients. An ethical permit was required and approved by the regional ethic committee in Stockholm 2017-01-18 (Diary number: 2016/2484-31/2).

## Results

At baseline, i.e. at time of thrombosis, the median age was 28 years (range 15-64 years) and there was a predominance of female patients (Table I). Three patients had hypercoagulability and the dominant arm was more commonly affected than the contralateral. Bilateral thrombosis was seen in 2 patients (2%).

In Table I baseline characteristics of the study population is presented. To be noted is that DASH-score baseline is not presented for the conservatively treated patient group, as the time point for baseline cannot be defined for this group.

Patients treated with early thrombolysis and surgery (acute group) had a lower DASH-score at long-term follow-up compared to the patients treated with delayed surgery (chronic group) p=.006 (ANOVA, F=5,5, df=2) (Table II). To be noted is that DASH baseline and DASH short-term for the conservative group is not presented, as the time points cannot be defined.

**Table II.** DASH-scores. DASH-score before and after intervention in relation to the different treatment groups.

|  | Acute (n=46) | Chronic (n=20) | Conservative (n=21) | P-value |
| --- | --- | --- | --- | --- |
| **DASH baseline (n)** | 32 | 13 |  | .080 |
| mean | 25.5 | 31.0 | ** |  |
| CI | 18.2-32.7 | 19.3-42.7 |  |  |
| **DASH short-term (n)** | 38 | 12 |  | .299 |
| mean | 11.2 | 18.9 | ** |  |
| CI | 7.2-15.2 | 10.4-27.4 |  |  |
| **DASH long-term (n)** | 33 | 14 | 18 | .006 |
| mean | 5.0 | 20.6 | 14.8 |  |
| CI | 1.8-8.2 | 9.4-31.8 | 4.2-25.5 |  |

Abbreviations: *DASH baseline*, DASH-score before intervention; *DASH short-term*, DASH-score up to 100 days after intervention; *DASH long term*, DASH-score longer than 100 days after intervention; CI, 95% confidence interval; \*\*, DASH baseline and DASH short-term can not be defined for this group

The long-term DASH-score was lower in the acute group compared to all patients not receiving early intervention i.e. chronic and conservative group together, p=.002 (ANOVA, F=10, df=1).

There was no indication of confounding by age, gender, smoking and affected arm (dominant/non dominant arm) in a multvariable regression model used (ANOVA, F=0.78, p=.55) i.e. the difference in long-term DASH-score between the groups could not be explained by these possible confounders.

The median long-term DASH-score for the three treatment groups in relation to the general DASH-score in a healthy population is illustrated in Figure II. As described above, DASH-score is age-dependent with higher DASH-scores the older the population (15). The mean DASH-score =2 in a normal population aged 19-34 years was used as a reference (34), as the median age of our study population was 28 years. (Figure II).

**Figure II.**
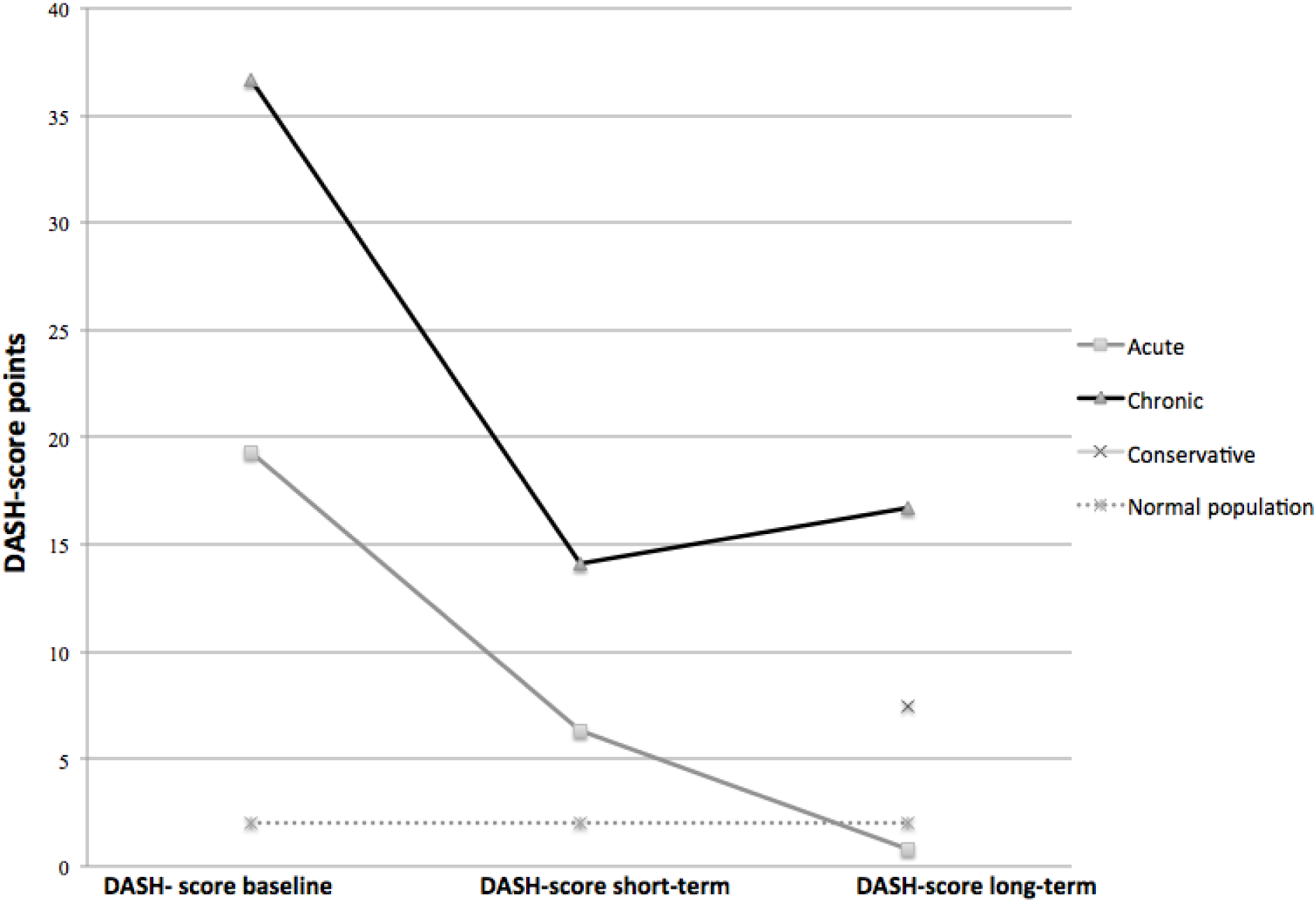
DASH-scores for the different treatment groups. Median DASH-score for the acute, chronic, conservative treatment group and normal population aged 19-34. DASH-score baseline and DASH-score short-term is not presented for the conservatively treated patient group, as the time point for baseline and short-term cannot be unambiguously defined for this group.

The mean follow-up time was 38 months (range 0-264 months). The proportion of patients with a clinically significant improvement (>13p decrease in DASH-score from baseline to long-term follow-up) was 62% (13/21) and 33% (3/9) in the acute and chronic group respectively.

## Discussion

This study aimed to investigate the associations between treatment strategy and QoL outcome in patients with PSVT. Our main finding was that QoL in patients who received early invasive treatment was significantly better than in patients who received delayed surgery, regardless of age, gender and other potential confounders. Furthermore, we had a larger proportion of female patients compared to other publications.

Studies comparing the three different treatment strategies for PSVT (anticoagulation alone, delayed decompressive surgery in patients with persisting symptoms after anticoagulation, and early invasive treatment with CDT and decompressive surgery), have shown conflicting results (21, 27, 29, 31).

Our results point to an improved QoL in patients treated with early invasive treatment, compared to a conservatively treated patient group. These results replicate to some extent the results from Taylor et al, using differences in DASH-score as a main outcome (29). They found a superior outcome in patients receiving early invasive treatment compared to patients treated with anticoagulation alone. Together, our results indicate that patients benefit from early intervention compared to either anticoagulation alone or delayed surgery. However, compared to our study, Taylor et al achieved an improvement in DASH-score for all surgically treated patients groups (including the patient group with delayed surgery) in relation to the group treated with anticoagulation alone. According to our protocol, conservatively treated patients received delayed surgery in case of persisting severe symptoms, consequently this group had the highest DASH-score at baseline (Table II).

Therefore, it can be argued that the delayed surgery group in our study contained patients with the most severe symptoms of all patients included in our study, which can further explain why they did not achieve a lower DASH-score at long-term follow-up than the conservatively treated patient group. Taylor et al used a shorter version of the DASH-form (Quick-DASH) than we did, which may have affected the precision in their study. Furthermore, the selection of patients for delayed surgery is not clearly described, making comparisons with our population difficult. The difference in DASH-score at baseline was smaller between the delayed surgery group and the group treated with anticoagulation alone compared to our study. Possibly, Taylor et al performed delayed surgery on patients with mild symptoms who may have improved without surgery. This could to some extent explain the difference between their and our results regarding the outcome in DASH-score long-term after delayed surgery.

To our knowledge, our study and the study performed by Taylor et al are the only studies evaluating QoL in patients with PSVT using the DASH instrument. Urschel et al published the first large material regarding PSVT, using an unstandardized evaluation of pain relief, return to work and limited recreation in order to interpret clinical results (21). Similar to our study, the best result was achieved in the patient group where early thrombolysis and surgery was performed. One limitation to this study was the lack of quality of life measurement, making it difficult to evaluate symptoms at baseline compared to symptoms at follow-up.

Furthermore, only 25 patients were treated with CDT and early surgery. However, the result from this study suggests early invasive treatment being superior to a conservative treatment.

Instead of describing relief of symptoms, some studies have reported on the vein patency after intervention (27, 31). Molina et al reported a 100% vein patency after early CDT and surgery (27), however an occluded vein is not necessarily equal to severe symptoms. Taylor et al had no difference in mean DASH-score at the final follow-up between patients who had an occluded subclavian vein or not (29). One theory discussed in the report by Taylor et al, is that if an occlusion of the vein occur, there might be a formation of additional collateral veins resulting in sufficient compensatory flow, which can give a symptom relief (29).

Even though studies have suggested that anticoagulation treatment often is insufficient to treat PSVT (20, 21), we had some patients treated with anticoagulation alone that achieved lower DASH-score than patients treated with delayed decompressive surgery. This suggests that some patients might be able to achieve good results from anticoagulation alone, and do not need surgery. Perhaps, this is because of the formation of collateral veins, as discussed above. The issue is that there is a lack of a marker for early identification of patients who will achieve good symptom relief with anticoagulation alone, and young and otherwise healthy individuals developing PSVT might not want to take the risk of developing PTS.

We compared the DASH-scores of our groups to the DASH-score of a general population within the same age-category as our study population, and found that the median long-term DASH-score was lower in the acute group compared to the general population. The conservative group had a DASH-score only a few points higher than the normal population, whilst the chronic group had the highest long-term DASH-score of all the groups. This indicates the importance of immediate diagnosis and treatment of PSVT-patients in order to reduce the risk of developing severe and chronic symptoms of the affected arm.

PSVT is usually referred to as a thrombosis affecting mostly young, athletic male, with a male to female ratio estimated to 2:1 (1, 6). Similar to other studies (29, 35), our study population had a larger proportion of women (54%). This might be explained by our centre not being a referral centre and therefore see a wide range of patients, not only male athletes, compared to what other big referral centres treating PSVT might do (21, 31). Lately it has been discussed if more women develop PSVT due to an increasing female athlete population, which can also be an explanation to why we have a larger portion of female in our study group (36).

Inherited thrombophilia increases the risk for developing PSVT (35). Compared to DVT within the lower extremities, it is less common with inherited thrombophilia for UEDVT (37). We report a frequency of 3% with a known clotting disorder in our study population, confirming a percentage similar to other studies (29, 31). To be noted is that patients within our study were not by default screened for inherited clotting disorders, because of its rarity it is not recommended with screening after PSVT in Stockholm. Patients were only screened for clotting disorders in case of present heritability, and therefore the true number of affected patients is not known.

### Strengths and limitations

Our study is one of the first studies to evaluate treatment for PSVT with a QoL instrument. This gives a better understanding of the disabilities due to the thrombosis compared to evaluation of vein patency only. Our clinic see a large proportion of detected patients with PSVT in county of Stockholm decreasing the risk for referral bias of the most symptomatic patients compared to tertiary referral centres. The follow-up time was in most patients long enough for developing PTS and the results are therefore mirroring the long-term results of the different treatment strategies. While other studies have used the abbreviated quick-DASH, we used the full DASH instrument, which may contribute to, increased precise QoL measurements.

This is a cohort study with the inherent risk of residual confounding, and therefore associations between treatment strategy and outcome could be due to other factors. Data was not crosschecked before analysis and therefore potential manual errors have not been accounted for. However, the data was entered in to the software EpiData, which was programmed to have only a few predetermined options for every variable entered in to the data-sheet. This decreased the risk of entering inadequate data in to the form before analysis. It can also be argued that potential manual errors would be random rather than systematic, and thereby not likely to change the outcome.

### Clinical applications

The best prevention of PTS after PSVT is today not known, and studies have provided inconsistent results. Nevertheless, the results of our study indicate that patients treated with early invasive treatment with CDT and surgery have a better outcome regarding QoL, compared to patients receiving delayed surgery. This stresses the importance of diagnosing and referring PSVT patients in time to have the option to offer early invasive treatment.

Furthermore, our results might indicate to have a lower threshold to perform immediate thrombolysis and surgery for this patient group.

### Future studies

More research is required in order to get some high-level evidence to provide the best treatment for PSVT patients. A randomized controlled study where PSVT patients are randomized to different treatment strategies is desired. However, this may be difficult to carry out because this is a young and otherwise healthy patient group and studies available point to better result if immediate treatment compared to conservative treatment. One possibility would be a design where all patients receive thrombolysis before randomization to either early surgery or anticoagulation for three to six months. This could answer if early surgery is necessary or can be performed in those who experience a rethrombosis after anticoagulation.

In our study, the group of patients that did not receive an early invasive treatment contained both patients with mild symptoms (conservative group) and patients experiencing severe symptoms who later had delayed surgery (chronic group). This suggests that not all patients require surgery but can achieve a good QoL with a treatment of anticoagulation alone. There is no marker that identifies patients who will benefit from anticoagulation alone and patients who will experience a reduced QoL on anticoagulation alone. This can be further explored in order to find a marker for identifying patients who could benefit from anticoagulation alone, not needing surgery.

## Conclusions

Treatment of PSVT with immediate thrombolysis and surgery was associated with superior QoL compared to treatment with delayed decompressive surgery. Because of the retrospective study design, associations between treatment strategy and outcome could be due to other factors. A randomized clinical trial comparing early versus delayed or no surgery is needed in order to provide high-level evidence regarding what treatment strategy is superior for PSVT patients.

## Abbreviations

CDT: Catheter-directed thrombolysis
CRF: Case report form
CT: Computed tomography
CVC: Central venous catheter
DASH: The disabilities of the arm, shoulder and hand
DVT: Deep venous thrombosis
EQ-5D: EuroQol five dimensions
MRI: Magnetic resonance imaging
PE: Pulmonary embolism
PSVT: Primary subclavian vein thrombosis
PTS: Post-thrombotic syndrome
QoL: Quality of life
UEDVT: Upper extremity deep venous thrombosis
VKA: Vitamin K antagonist
V-TOS: Venous thoracic outlet syndrome

## Data Availability

Data will be shared for individual-participant-data meta-analysis with other members of the research community who have an affiliation to a recognised medical university. Data will only be shared with investigator support, after approval of a proposal, and with a signed data access agreement. Additional restrictions will apply in accordance with Swedish law.

